# LLM-based Multi-Agent Collaboration for Abstract Screening towards Automated Systematic Reviews

**DOI:** 10.1101/2025.08.11.25333429

**Authors:** Opeoluwa Akinseloyin, Xiaorui Jiang, Vasile Palade

## Abstract

**Objective:** Systematic reviews (SRs) are essential for evidence-based practice but remain labor-intensive, especially during abstract screening. This study evaluates whether multiple large language model (multi-LLM) collaboration can improve the efficiency and reduce costs for abstract screening.

**Methods:** Abstract screening was framed as a question-answering (QA) task using cost-effective LLMs. Three multi-LLM collaboration strategies were evaluated, including majority voting by averaging opinions of peers, multi-agent debate (MAD) for answer refinement, and LLM-based adjudication against answers of individual QA baselines. These strategies were evaluated on 28 SRs of the CLEF eHealth 2019 Technology-Assisted Review benchmark using standard performance metrics such as Mean Average Precision (MAP) and Work Saved over Sampling at 95% recall (WSS@95%).

**Results:** Multi-LLM collaboration significantly outperformed QA baselines. Majority voting was overall the best strategy, achieving the highest MAP 0.462 and 0.341 on subsets of SRs about clinical intervention and diagnostic technology assessment, respectively, with WSS@95% 0.606 and 0.680, enabling in theory up to 68% workload reduction at 95% recall of all relevant studies. MAD improved weaker models most. Our own adjudicator-as-a-ranker method was the second strongest approach, surpassing adjudicator-as-a-judge, but at a significantly higher cost than majority voting and debating.

**Conclusion:** Multi-LLM collaboration substantially improves abstract screening efficiency, and the success lies in model diversity. Making the best use of diversity, majority voting stands out in terms of both excellent performance and low cost compared to adjudication. Despite context-dependent gains and diminishing model diversity, MAD is still a cost-effective strategy and a potential direction of further research.

## Background and Significance

Systematic reviews serve as cornerstones for evidence-based practice, particularly in medicine and healthcare, by providing rigorous summaries of existing knowledge [1]. However, conducting SRs is notoriously labor-intensive, often requiring researchers to spend over several months [2, 3]. A primary bottleneck is the title and abstract screening phase, where the sheer volume of retrieved studies—sometimes as high as tens of thousands [4, 5]—presents formidable challenges, amplified by the standard practice of involving multiple human annotators [6].

There has been two decades of efforts on using AI to automated or semi-automate the screening task since Cohen et al.’s seminal works [7, 8]. Early automation efforts leveraged machine learning techniques [2, 9], later progressing to deep learning models [10, 11]. These approaches faced significant challenges, such as the requirement for extensive labelled data, extreme class imbalance and the need for model retraining for each new review [2, 12, 13, 14, 15]. Active learning sought to alleviate these issues through iterative human feedback [16, 17, 18, 19], but faced challenges in achieving a high degree of automation due to the zero-shot nature of this problem [20, 21].

The advent of Large Language Models (LLMs) marked a paradigm shift, offering unprecedented zero-shot learning capabilities for tasks like abstract screening [22, 23, 24, 25, 26, 27]. Recent studies have demonstrated LLMs’ potential across various SR stages, from search query generation [28] to literature screening tasks [29, 30, 31, 32] and even the whole SR pipeline [33]. As shown in [19] and [34], LLMs can also make the active learning process for abstract screening more efficient. However, single LLMs inevitably suffer inherent model biases [35, 36] and weak alignment with nuanced human judgment [37], making it more less likely for individual LLMs to meet the sensitivity/recall requirement and reach a good balance between sensitivity and specificity [38]. These limitations have catalyzed interest in enhancing reliability through multiple large language model (multi-LLM) collaboration, aka LLM-based Multi-Agent Systems (MAS) [39, 40, 41]. For example, ensemble approaches, a primitive form of multi-model collaboration, have demonstrated superior performance for abstract screening by combining the decisions from a range of LLMs [38, 42, 63] to achieve higher sensitivity/recall and better balance between sensitivity and specificity/precision.

LLM-based MAS (here agent is an LLM) has emerged as a powerful paradigm across various domains, with extensive research demonstrating their effectiveness [39, 44, 45]. Key collaboration strategies include debating mechanisms [46, 47], where agents engage in structured argumentation [46, 48, 49], and adjudication approaches leveraging, for example, the LLM-as-a-Judge frameworks [50, 51]. These approaches aim to mitigate individual model weaknesses through collective reasoning among peers and have been demonstrated effective for complex reasoning tasks in medical domains [52, 53, 54]. Yet, a comprehensive investigation of multi-LLM collaboration hasn’t been presented in the context of abstract screening. This paper embarks on the first comprehensive investigation into multi-LLM collaboration in automated abstract screening. Our contributions are threefold: (1) We investigate multi-LLM collaborative strategies, including ensembling, debating, and adjudication, towards systematic review automation. (2) We comparatively evaluate different strategies to identify the most robust approaches. (3) We empirically analyze the core success factors that enable multi-LLM collaboration to mitigate errors of individual LLMs.

## Methodology

### LLM-based Question Answering for Screening

Consistent with [30], we formulate the abstract screening task using a question-answering (QA) framework. Suppose each SR constitutes an unannoated dataset of (the titles and abstracts of) candidate studies (i.e., documents) *D* = {*d*_1_, *d*_2_, ⋯, *d*_*N*_}, where N is the total number of documents and di is the *i*-th document. Abstract screening is the process of assigning a label to indicate that a document should be “included” into or “excluded” from the remaining steps of an SR, for which screening prioritization ranks the documents in descending order of their likelihood of being included.

Each SR has a paragraph of selection criteria questions, *Q* = {*q*_1_, ⋯, *q*_*K*_ }, that every included study must satisfy. Given a document *d*_*i*_ (*i* = 1, ⋯, *N*), each inclusion criteria question *q*_*k*_ (*k* = 1, ⋯, *K*) will be answered by an LLM-based QA model *M*^qa^, with the answer *a*_*i,k*_^*M*qa^ being either “Positive” (meaning meeting the criterion), “Negative” (meaning not) or “Neutral” (meaning unsure or not answerable) plus a reasoning text. To ease discussion, the QA process is formalized as follows, with the prompt and a corresponding example presented in Appendix A and B of Supplementary File 1 respectively:

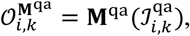

where the input ℐ_*i,k*_^qa^ = 〈*q*_*k*_, *di*〉 contains the *k*-th inclusion criteria question on the *i*-th document, and the output *O*_*i,k*_^*M*qa^ = 〈*a*_*i,k*_^*M*qa^, *r*_*i,k*_^*M*qa^ 〉 contains the answer *a*_*i,k*_^*M*qa^ and its reasoning *r*_*i,k*_^*M*qa^ for the *k*-th question on the *i*-th document.

Given a QA model *M*^qa^, we use the same method in [30] to score *d*_*i*_ with respect to each question *q*_*k*_, by assigning the probability that the corresponding answer text (i.e., the concatenation of *a*_*i,k*_^*M*qa^ and *r*_*i,k*_^*M*qa^ has a positive sentiment, according to a pretrained BART model [55]:

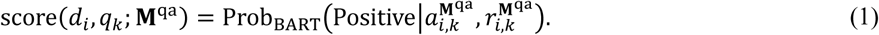

The document score is the sum of its scores with respect to all inclusion criteria:

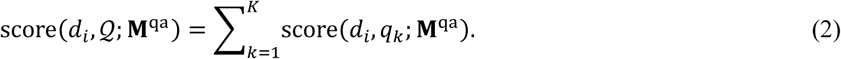

### LLM-based Question Answering for Screening

Consistent with [30], we formulate the abstract screening task using a question-answering (QA) framework. Suppose each SR constitutes an unannoated dataset Inspired by recent work in LLM-based MAS [39, 46, 56], we investigate three distinct strategies for combining the outputs and reasoning from multiple LLMs (aka agents). The prompts for all strategies are presented in Appendix A in Supplementary File 1, while Appendix B presents an illustrative example for each of the three strategies.

### Majority Voting

Assuming the “wisdom of the crowd” supersedes individuals, the first simple but extremely effective strategy is majority voting, more precisely soft voting (Soft-Vote). Given a collection of primary (QA) models ℳ = {*M*_*l*_}|^*L*^_*l*=1_, for each question *q*_*k*_and document di, the soft voting score is defined as the average of the scores for each primary model:

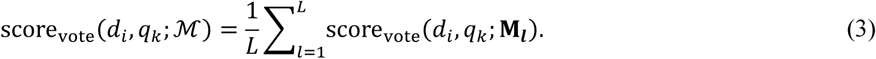

Then the final score for each document is the sum of the soft voting scores with respect to all inclusion criteria:

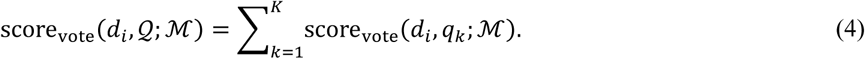

### Multi-Agent Debate

This strategy introduces a step of cross-agent communication, reflection, and refinement [46, 57]. After an initial round of independent QA, each agent is presented with the answers and reasoning of other agents for the same question on an abstract, and is prompted to reconsider its initial answer and reasoning in light of the perspectives of its peers. If an answer is changed, the updated answer and reasoning are recorded.

Formally, the debating process is defined as follows:

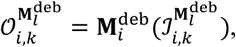

where *M*_*i*_^deb^ ∈ ℳ (l = 1, ⋯, L) is the debating model, the input ℐ_*i,k*_^*M*deb^ is the union of the output of itself and the outputs of all other QA models, plus the original QA input, i.e.,

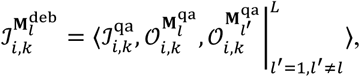

and the output of the debating model is as follows

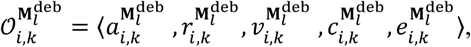

with a_*i,k*_^*Ml*deb^, r_*i,k*_^*Ml*deb^ and *e*_*i,k*_^*Mi*deb^ being the new ***a***nswer, the ***r***easoning, and the confidence ***v***alue of the debating agent on the new answer, *c*_*i,k*_^deb^ indicating whether the debating agent ***c***hanges the original answer (“Yes” or “No”), and *e*_*i,k*_^deb^ holding the ***e***xplanation for why the original answer is changed or kept unchanged Note that *M*^deb^ and *M*^qa^ refer to the same model *M*_*l*_ ∈ ℳ used for different purposes, taking different inputs and generating different outputs. Note that the new reasoning *r*_,*k*_^deb^ is the updated rationale for the current answer, while the explanation *e*_,*k*_^deb^ explicitly states why the answer was changed or kept unchanged after considering peers’ input. See Appendix C and D in Supplementary File 1 for an example and further explanations.

The final scoring and ranking are based on the answers after debating. Formally, given a collection of LLMs, amongst which *M*^deb^ is the debating model, the debating score for each document is defined as follows:

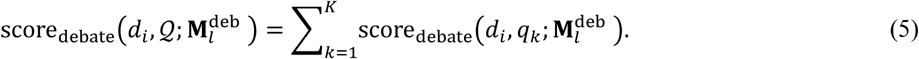

where

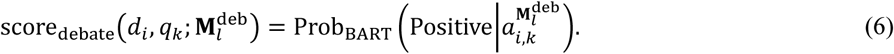

Note that here the new reasoning *r*_*i,k*_^*l*^ is not used for scoring because the debating model may change the answer and the explanation for such change may contain negative information against the old answer rather than negative viewpoints against the question, in which case the linguistic cues for answer justification will mislead BART in score assignment. Instead, scoring is based solely on the final answer *a*_*i,k*_^*l*^. In cases where documents receive identical scores, the debating agent’s confidence *e*_*i,k*_^*l*^ is used to break ties by producing a weighted score, ensuring that answers supported with higher certainty are prioritized in the final ranking. Also note that our MAD approach, together with LLM-based adjudication to be introduced below, can be seen as variants of the idea of exchange-of-thought [48].

### LLM-based Adjudication

The third strategy uses a separate and more powerful LLM as the adjudicator to synthesize different opinions and make the final verdict [58]. For each question, the adjudicator receives the initial answers of all primary models, analyzes their reasoning texts, and determines which answer is the most accurate or well-justified. Formally, given a collection of LLMs ℳ as QA models, for each inclusion criteria question *q_k_* on a document *d_i_*, the judging process of the adjudicator LLM *M*^adj^ ∉ ℳ is defined as follows:

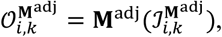

where the input ℐ_*i,k*_^*M*adj^ is exactly the same as ℐ_*i,k*_^*M*deb^ and the output is defined as follows,

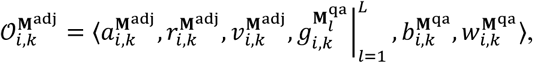

with *a*_*i,k*_, *r*_*i,k*_, and *e*_*i,k*_ being the adjudicator’s new ***a***nswer, its ***r***easoning and confidence ***v***alue, each *g*_*i,k*_^*l*^ being the grading of a QA model (a rating value between 0 and 1), and *b*_*i,k*_^*M*qa^ and *w*_*i,k*_^*M*qa^ being the best and worst models selected by the adjudicator, respectively.

Two variants are proposed.

### Adjudicator as a Judge

The first variant is to use this LLM adjudicator as a “Judge” [50], or called “meta-agent” in other literature [47]. Similar to other methods, the score of a document di with respect to each criteria question *q_k_* by an adjudicator as a judge is the probability that its answer text (i.e., *a*_*i,k*_^*M*adj^ + *r*_*i,k*_^*M*adj^) has a positive sentiment based on BART:

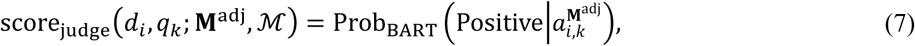

and the final score of each document is:

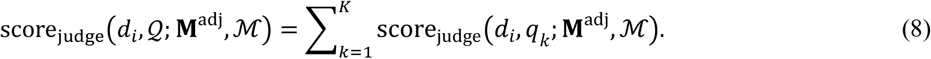

### Adjudicator-as-a-Ranker

Alternatively, a separate LLM (called adjudicator) is asked to rate the quality of individual answers and generate a grade for each primary model, denoted by *g*_*i,k*_^*l*^ (*l* = 1, ⋯, *L*), which are then used to calculate a weighted average of the primary models’ scores, i.e., the score of a document *d_i_* with respect to criteria question *q_k_* by the adjudicator:

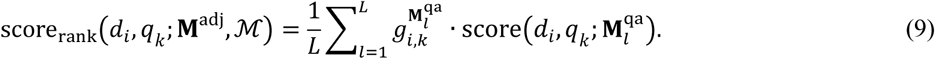

Then, the final score for each document according to the adjudicator as a ranker is:

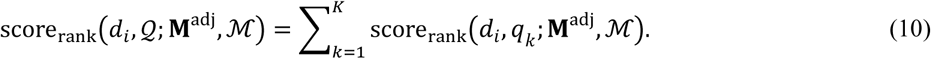

### Re-ranking

Screening prioritization performance can be further improved through re-ranking as in [30]. The rationale is an included study should meet all selection criteria or most of them (when there are certain criteria unanswerable), so we can expect a high semantic relevance between the requirements of an SR’s inclusion criteria and the information in an included study.

#### Macro-level Re-ranking (rr-mac)

Relevance is measured by the cosine similarity between the text embeddings of each candidate study and the selection criteria paragraph, denoted by rel(*d*_*i*_, *Q*). A macro-level re-ranking score is calculated as follows:

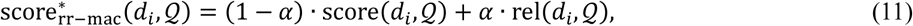

where α ∈ (0, 1), and score(*d*_*i*_, *Q*) is the score of *d_i_*that is calculated according to either Eq. (4), Eq. (5), Eq. (8) or Eq. (10).

#### Micro-level Re-ranking (rr-mic)

Cosine similarity is calculated between each included study and each inclusion criterion *q*_*k*_ ∈ *Q*, denoted by rel(*d*_*i*_, *q*_*k*_). Micro-level re-ranking first calculates a new score for each document with respect to each question as follows:

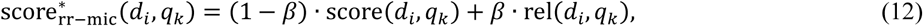

where β ∈ (0, 1) and score(*d*_*i*_, *q*_*k*_) is calculated according to either Eq. (3), Eq. (6), Eq. (7) or Eq. (9). Then, a new score for each document is calculated by summing up the scores with respect to all criteria questions:

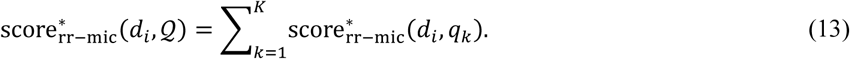

### Experimental Setup

#### Dataset and Evaluation Metrics

Evaluation is done on CLEF eHealth 2019 Task 2: Technology-Assisted Reviews in Empirical Medicine (TAR2019)—a famous standard benchmark for evaluating abstract screening methods, including 20 Cochrane reviews about clinical intervention (Intervention; in total 39,792 documents) and 8 reviews about diagnostic technology assessment (DTA; 26,830). For all experiments in this study, each document’s title was concatenated with its abstract before being passed to the LLM. This ensured that inclusion cues present in titles such as study design, population characteristics, or intervention types were captured alongside the more detailed information typically found in abstracts. For the small subset of records in TAR2019 that contained only titles without abstracts, we processed these using the title text alone. Across all reviews, 66,622 documents were screened, with inclusion rates ranging from 0.2% to 36.1% (mean: 5.8%). Detailed per-review statistics, including exact document counts, inclusion rates, selection criteria paragraphs, and generated question sets for each review, are provided in Appendix D of Supplementary File 1.

We employ standard TAR evaluation metrics, including the rank of the *L*ast *Rel*evant document (*L*_Rel_), *M*ean *A*verage *P*recision (MAP), *R*ecall at top *k*% of documents screened (*R*@*k*%, for *k* = 5, 10, 20, …, 50), and *W*ork *S*aved over *S*ampling (WSS) at the recall level of *R*% (WSS@*R*%, for *R* = 95, 100) [7]. Metrics are calculated per SR and then averaged across all the SRs in each of the two categories.

### Large Language Models and Baselines

LLM selection is based on two factors: balance between capability and computational cost, and popularity of model family. In the experiments, the “lightweight” versions of the GPT, Gemini and Claude families are chosen:

- GPT-4o Mini (gpt-4o-mini-2024-07-18),
- Claude 3 Haiku (claude-3-haiku-20240307), and
- Gemini 1.5 Flash (gemini-1.5-flash-preview-0514).

The LLM adjudicator, acting as both the “Judge” agent and the “Ranker” agent, is:

- Gemini 1.5 Pro (gemini-1.5-pro-preview-0514).

To ensure reproducibility, the temperature parameters for all models are set to zero. The models for evaluation include:

- Three primary QA models according to Eq. (2), named by `GPT`, `Haiku`, and `Gemini`;
- The soft voter according to Eq. (4), named by `Soft-Vote`;
- The three MAD models according to Eq. (5)), named by `GPT-MAD`, `Haiku-MAD`, and `Gemini-MAD`;
- The adjudicator-as-a-judge (`Adj-Judge`) and adjudicator-as-a-ranker (`Adj-Rank`) methods according to Eq. (8) and Eq. (10) respectively.

We also compare the re-ranking variants of the aforementioned approaches that integrate both macro- and micro-level re-ranking, according to Eqs. (11) and (13). The GPT embedding model “text-embedding-ada-002” is used to generate text embeddings for relevance calculation.

To ensure reproducibility, all prompts used in this study are provided in Appendix A of Supplementary File 1. All experiments used a temperature parameter of 0.1 to maximize reproducibility and deterministic outputs ^1^. Implementation code, model parameters, and evaluation scripts are available at the following link under the MIT License: https://github.com/Ope-Akinseloyin/Multi_LLM-Citation-Screening.git.

## Results and Discussion

### Main Results: Screening Performance Enhancement

Table 1 presents the performance comparisons, which are summarised in the following subsections.

**Table 1.**
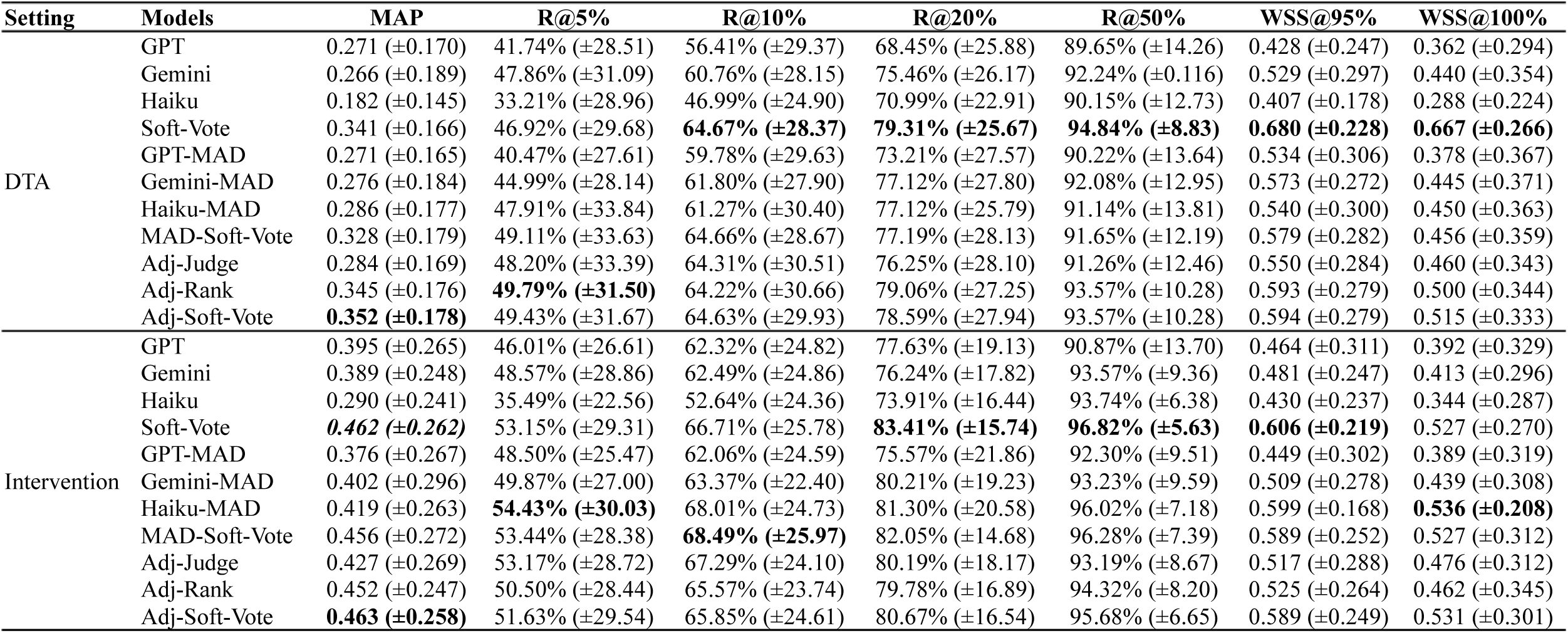
Performances of Multi-LLM Collaboration.

### Majority Voting

On both DTA and Intervention, Soft-Vote substantially outperformed all individual QA baselines with statistically significant improvements in MAP (paired *t*-test: *p* < 0.001 for all pairwise comparisons; Wilcoxon signed-rank test: *p* < 0.001) and WSS@95% (*p* < 0.01 for all comparisons), obtaining the highest MAP, recall and WSS values among all competitors. Mean R@10% reached 64.67% (±28.37) and 66.71% (±25.78) on DTA and Intervention respectively, which demonstrated this LLM ensemble’s strong ranking capability for abstract screening. In term of theoretical workload savings, it achieved 0.680 (±0.228) WSS@95% and 0.667 (±0.266) WSS@100% on DTA, and 0.606% (±0.219) WSS@95% and 0.527 (±0.270) WSS100%. Considering its low cost, the simple approach `Soft-Vote` is both strong and cost-effective (refer to Appendix E in Supplementary File 1 for the detailed breakdown for human cost estimation). The simple aggregation effectively mitigates individual model weaknesses and biases. The theoretical average workload reductions can be as high as approximately 68.0% and 60.6% on the 8 DTA and 20 Intervention SRs, respectively when the recall threshold of relevant studies is 95%, satisfying a widely-adopted critical requirement for avoiding causing significant damage to the reliability of the conclusions of an SR.

### Multi-Agent Debate

The debating strategy showed model-specific benefits. For most primary QA models in both the DTA and Intervention settings, MAD improved over QA baselines on most performance metrics except for `GPT-MAD`. `GPT-MAD`’s performances on DTA slightly dropped in terms of MAP and R@5%, while significantly outperforming GPT in term of WSS. On Intervention, `GPT-MAD` had more mixed performances. It recorded a slight drop in MAP and WSS, but performed slightly better on recall. Comparatively, `Gemini-MAD`’s performance gain over its QA counterpart is much more stable. Across DTA and Intervention, `Gemini-MAD` outperformed `Gemini` on most performance metrics such as MAP, WSS@95% and WSS@100%. Although the performance gains of `Gemini-MAD` cannot be said significant, they are not marginal either. Also note that, `Gemini` is the strongest QA model, outperforming other QA baselines by large margins.

Notably and surprisingly, it was the weakest model `Haiku` which benefited most from MAD. On DTA, H`aiku-MAD`’s MAP and WSS@95% increased from 0.182 and 0.407 to 0.286 and 0.540, respectively, while on Intervention from 0.290 and 0.430 to 0.419 and 0.599. It is worth noting that, on Intervention, `Haiku-MAD` was overall the second best-performing model and was the best in term of R@5% and WSS@100%. In summary, the debating models, except GPT-MAD, exhibited good performances almost on par with the much more expensive adjudication methods, which highlights a promising future of MAD systems in systematic review automation.

### LLM-based Adjudication

Between LLM-based adjudication approaches, the adjudicator-as-a-ranker method (`Adj-Rank`) outperformed adjudicator-as-a-judge (`Adj-Judge`) in most important metrics like MAP, R@50% and WSS@95%, demonstrating that *preserving diverse perspectives through weighted averaging* is likely more effective than selecting a single “best” answer. Although adjudication did not beat `Soft-Vote`, it was much stronger than most other competitors in both settings except that both adjudicator variants were outperformed by `Haiku-MAD` on Intervention. These results not only underscore the potential value of the increasingly popular “LLM-as-a-Judge” paradigm [50] and the superiority of adjudicator-as-a-ranker as a novel contribution to this paradigm, but also highlight the high potential of the multi-agent debating paradigm, especially when cost-effectiveness is considered.

### Per-Review Investigation

Individual review performance varied substantially. Comprehensive per-review breakdowns showing all metrics for each systematic review are provided in Supplementary Files 2-3, enabling readers to assess method performance under conditions similar to their specific review characteristics. Multi-LLM collaboration showed greater benefits for reviews with moderate inclusion rates (5–15%) and higher inter-model disagreement, as evidenced by the ablation study in Figure 1 and the correlation analysis in Figure 2. `Soft-Vote` achieved consistent improvements across all 28 reviews (MAP range: 0.182–0.687.), while benefits from multi-agent debate were most pronounced for weaker models (`Haiku-MAD`: +0.129 MAP improvement on Intervention, *p* < 0.001). The bootstrap confidence intervals of the performances of each method are presented in Supplementary File 4. Comprehensive statistical comparisons, including effect sizes and bootstrap confidence intervals, are provided in the Supplementary File 5.

**Fig. 1.**
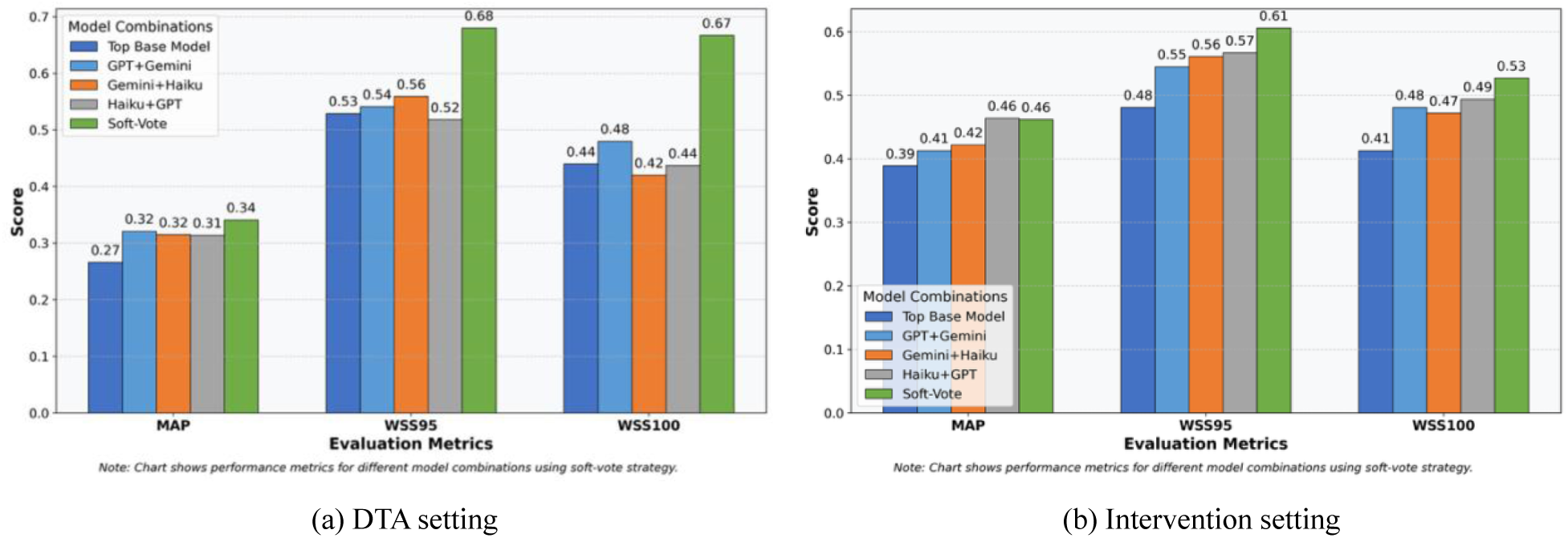
Ablation Study of Majority Voting.

**Fig. 2.**
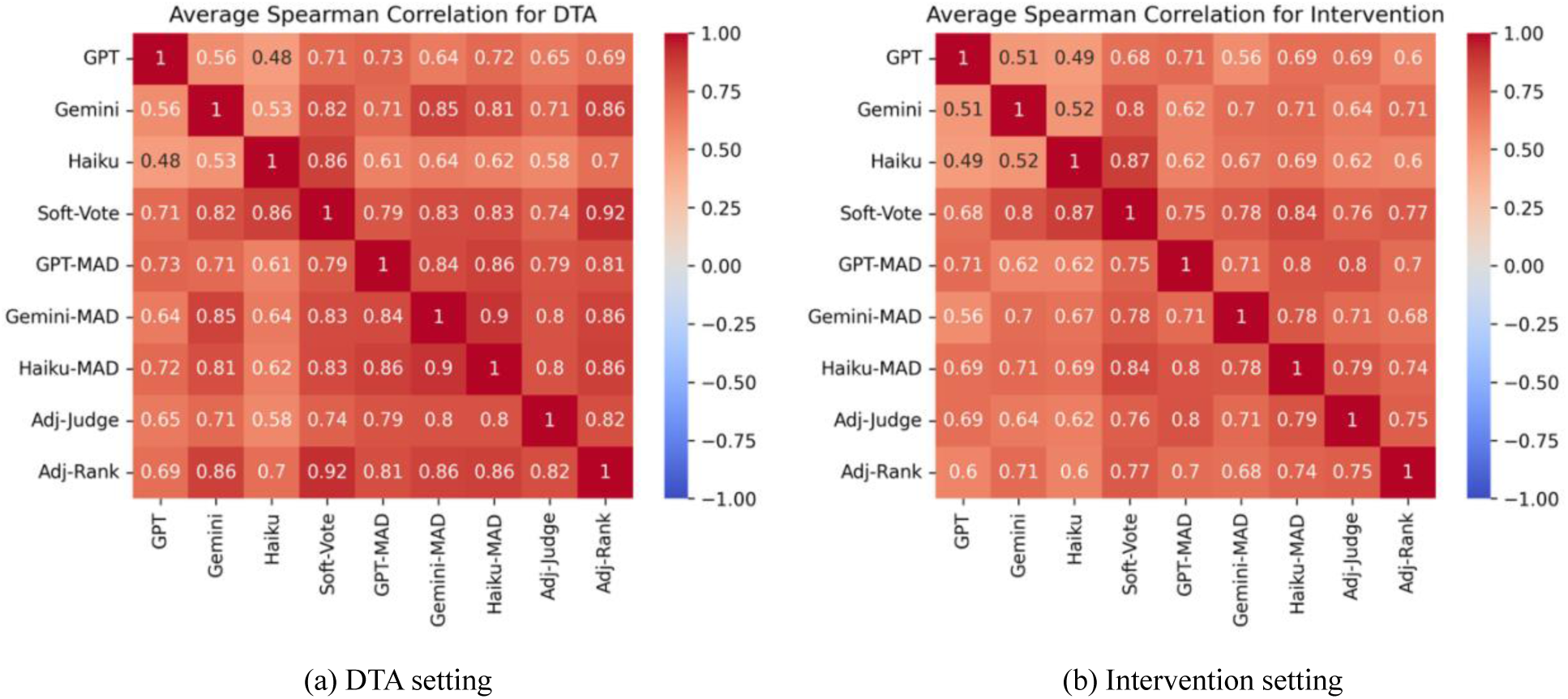
Correlation Analysis Between Models.

### Insights, Observed Strengths and Weaknesses

#### Critics of Majority Voting

The majority voting strategy `Soft-Vote` demonstrates consistent superiority (Table 1). Figure 1 shows the results of ablation study of this simple ensemble approach where we see that any two-model combination also achieved substantial improvements over individual models. Particularly, `GPT`+`Haiku` performed surprisingly well, almost rivalling `Soft-Vote` on Intervention in some metrics, like MAP (0.46 vs. 0.46), WSS@95% (0.57 vs. 0.61), and WSS@100% (0.49 vs. 0.53). These findings suggest that benefits from diversity begin with just two base models and increase as more models are added.

Correlation analysis in Figure 2 reveals moderate correlations between the QA models (Spearman’s Rank Correlations: 0.48–0.56 on DTA and 0.49–0.52 on Intervention), indicating each primary model captures different aspects of relevance—a diversity that an ensemble approach can effectively leverage. `Soft-Vote` shows high correlation with each individual model (Spearman: 0.71–0.86 on DTA and 0.68–0.87 on Intervention), suggesting it preserves their strengths while mitigating weaknesses. The central role of diversity will be discussed in more detail in the “Model Diversity” subsection.

The strong performance combined with computational efficiency (see the cost breakdown in Table 2 and Appendix E in Supplementary File 1 for the detailed cost estimation), which is less than 1/14 of adjudication and at most around 1/186 of the cost of a single human reviewer. This positions Majority Voting as an excellent default choice for early-stage screening. The ablation results suggest that even resource-constrained implementations using just two diverse models can achieve significant benefits.

**Table 2.** LLM Pricing, Cost Breakdown, and Runtime.

| Setting | Type | Model | Input Price* | Input Cost | Output Price* | Output Cost | Total Cost | Time (h) |
| --- | --- | --- | --- | --- | --- | --- | --- | --- |
| DTA | Question Answering | GPT-4o Mini | 0.15 | \$2.54 | 0.6 | \$8.54 | \$11.08 | 100.6 |
| | | Gemini 1.5 Flash | 0.075 | \$1.27 | 0.3 | \$4.17 | \$5.44 | 39.32 |
| | | Claude 3 Haiku | 0.25 | \$4.24 | 1.25 | \$23.66 | \$27.90 | 62.5 |
| | Majority Voting | Soft-Vote | -- | \$8.05 | -- | \$36.38 | \$44.43 | 202.42 |
| | Debating | GPT-4o Mini | 0.15 | \$18.32 | 0.6 | \$47.38 | \$65.70 | 334.52 |
| | | Gemini 1.5 Flash | 0.075 | \$13.19 | 0.3 | \$42.64 | \$55.82 | 252.3 |
| | | Claude 3 Haiku | 0.25 | \$25.16 | 1.25 | \$65.15 | \$90.31 | 280.26 |
| | Voting on debating | MAD-Soft-Vote | -- | \$56.67 | -- | \$155.17 | \$211.84 | 462.24 |
| | Adjudication | Gemini 1.5 Pro | 3.5 | \$263.49 | 10.5 | \$389.65 | \$653.15 | 906.32 |
| Intervention | Question Answering | GPT-4o Mini | 0.15 | \$4.25 | 0.6 | \$12.56 | \$16.81 | 149.0 |
| | | Gemini 1.5 Flash | 0.075 | \$2.12 | 0.3 | \$5.55 | \$7.67 | 58.27 |
| | | Claude 3 Haiku | 0.25 | \$7.08 | 1.25 | \$34.65 | \$41.73 | 92.6 |
| | Majority Voting | Soft-Vote | -- | \$13.45 | -- | \$52.76 | \$66.21 | 299.8 |
| | Debating | GPT-4o Mini | 0.15 | \$28.77 | 0.6 | \$68.74 | \$97.51 | 495.97 |
| | | Gemini 1.5 Flash | 0.075 | \$21.11 | 0.3 | \$61.76 | \$82.88 | 373.92 |
| | | Claude 3 Haiku | 0.25 | \$38.98 | 1.25 | \$94.45 | \$133.43 | 415.22 |
| | Voting on debating | MAD-Soft-Vote | -- | \$88.86 | -- | \$224.95 | \$313.81 | 685.3 |
| | Adjudication | Gemini 1.5 Pro | 3.5 | \$394.11 | 10.5 | \$565.80 | \$959.90 | 1342.8 |

\* Input and output prices are expressed in dollars per million tokens.

#### Critics of Multi-Agent Debate

The debating strategy aims to improve screening performance by leveraging collective intelligence through exchanging structured argument. Overall, the MAD results provide several useful insights for designing MAD systems. Multi-agent debate indeed brings performance gains, even with agents that are much weaker. The rethinking process is obviously the key to the success of MAD, which arguably improves performance through three mechanisms: (1) error correction when agents encounter opposing viewpoints with supporting evidence, (2) uncertainty reduction by considering multiple perspectives on ambiguous cases, and (3) mitigation of model-specific bias through exposure to alternative interpretations.

Meanwhile, our analysis shows model-dependent outcomes as in Figure 3. Benefits brought by MAD seem to be affected by agents’ capabilities. While weaker agents will likely bring less benefits to stronger ones, they may benefit more from exposure to alternative perspectives. In reverse, opinions from stronger peers may have more significant impacts on self-reflection and decision-making. Both conform well with common sense. More specifically, `Gemini-MAD` showed minimal to negligible improvement over the QA baseline, possibly due to the latter’s high performance, less influence from external arguments, or suboptimal prompt tailoring. `GPT-MAD` exhibited more varied results. Despite obvious improvements on DTA, `GPT-MAD` stumbled at improving screening performance on Intervention across various metrics. Comparatively, `Haiku-MAD` obtained pronounce gains from debating. Although `Haiku` is notably much weaker than the other two competitors in medical abstract screening, it demonstrates a clear capacity for reasoning refinement through interaction with peers. This phenomenon may have shed some *unusual light on the design of cost-effective LLM-based MAD systems* because stronger individual models may not always benefit most from multi-agent debate.

**Fig. 3.**
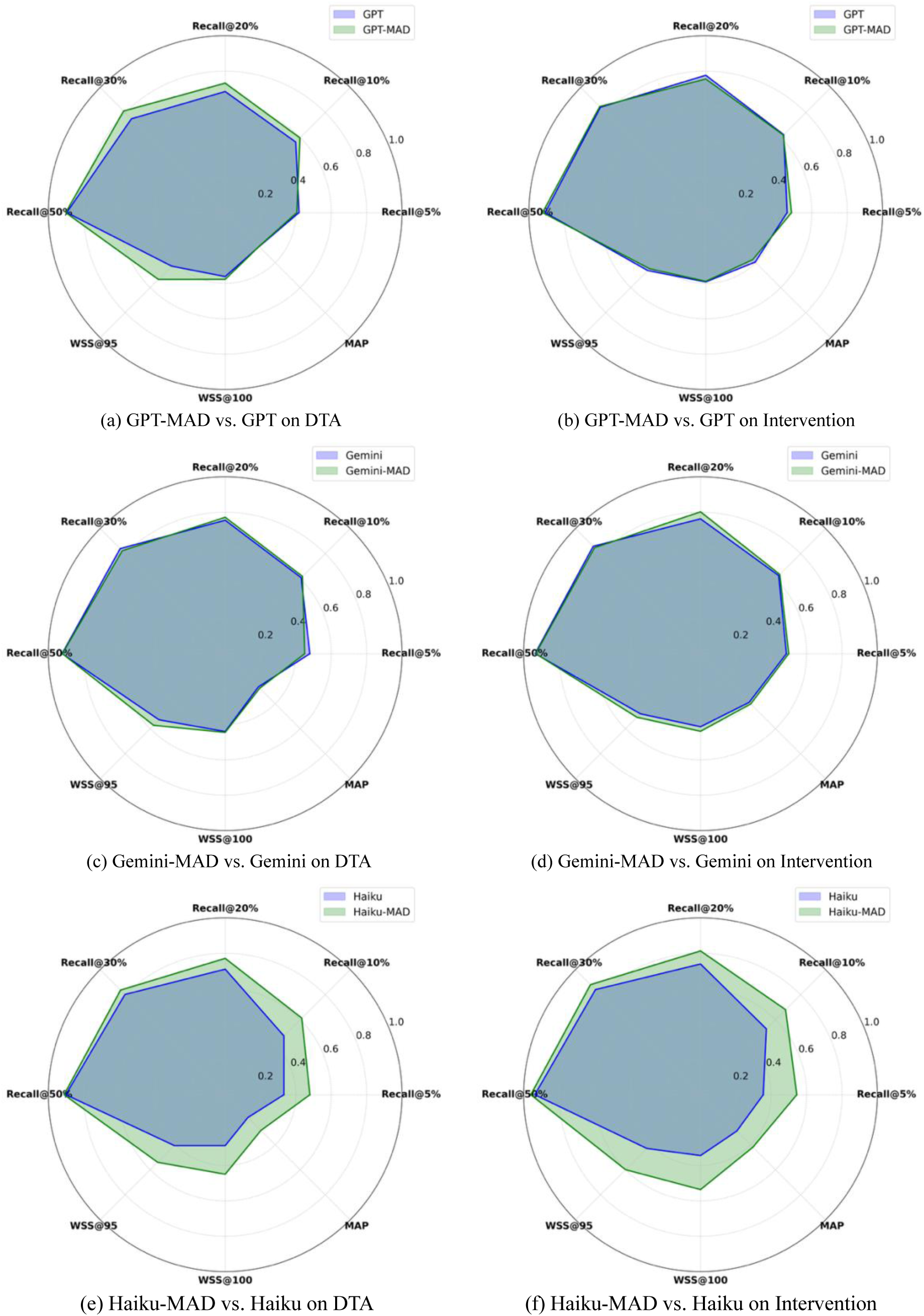
Multi-Agent Debate vs. QA Models

The success of `Haiku-MAD` may to some extent lie in `Haiku` being more “diverse” from `GPT` (seen from Haiku’s comparatively low correlations with others: for example 0.48 and 0.53 on DTA in Figure 2), but should be more rooted in its capability of leveraging peers’ wisdom. For example, although Gemini 1.5 Flash is the strongest individual model, it inclines to stick to its own decision, which can be demonstrated by the high correlation between the debating model `Gemini-MAD` and the QA baseline `Gemini` (e.g., 0.85 on DTA and 0.7 on Intervention). GPT-4o Mini has the same but slightly weaker inclination. For instance, `GPT-MAD` is most correlated with `GPT` at 0.73 on DTA and 0.71 on Intervention. Notably and surprisingly, Claude 3 Haiku is the only debating model which perhaps takes more peer opinions than its own, which is implied from the fact that `Haiku-MAD` is more correlated with `GPT` and `Gemini` than with `Haiku`. The findings may have an important implication: *It might not be individual agent’s own capability but its capability of assimilating different opinions that makes multi-agent debate systems work*.

The experiments also raise a question about whether MAD maintains model heterogeneity. Correlations among the three debating models have become much stronger than the correlations among the QA models, which is an expected phenomenon when peers gradually converge with the cohort while the cohort collectively improve. The ensemble over debating models, `MAD-Soft-Vote`, does not improve a lot over the best debating models (see Table 1), which further demonstrates the *core role of model heterogeneity* [59].

#### Critics of Adjudication

Adjudication strategies introduce a hierarchical decision-making layer in a multi-LLM collaborative framework. Table 3 shows the average score assigned to each primary model by the adjudicator (here Gemini 1.5 Pro), along with how often each primary model was deemed the best or worst performing model by the adjudicator. The “Main Results: Screening Performance Enhancement” subsection shows `Gemini` (Gemini 1.5 Flash) is the strongest individual model. Indeed, the Gemini 1.5 Pro adjudicator has selected `Gemini` (Gemini 1.5 Flash) as the “best model” in over 50% and 45% of cases of DTA and Intervention, respectively, while its opinions about `GPT` (GPT-4o Mini) and `Haiku` (Claude 3 Haiku) were more or less balanced. Not surprisingly, `Haiku` was most frequently deemed the “worst model”. On Intervention, the chance of `Haiku` being rated as the worst model is significantly higher, which is likely linked to the fact that Haiku’s performance gap from `GPT` is also bigger. On the contrary, the chance of `Gemini` being rated as the best model drops possibly because `Gemini`’s performance gain over other models on Intervention is less significant than on DTA. These findings have several implications. Firstly, the adjudicator may indeed have some good capabilities in deciding the more appropriate primary model on a case by case basis, a key reason for significant screening performance improvement by both adjudication methods. In the meanwhile, they have also revealed *a potential bias of the adjudicator towards its own model family*, in corroboration with findings reported in [60]. This is also reflected by the fact that the correlations between `Gemini` and the adjudicators is often stronger (Figure 2), although more experiments are expected in the abstract screening context. This potential bias is a critical consideration apart from the significantly higher cost, as it could inadvertently reinforce existing preferential bias or limit the diversity of perspectives.

**Table 3.** Ratings of QA Models by the Adjudicator.

| Setting | Model | Avg Rating | Best (%) | Worst (%) |
| --- | --- | --- | --- | --- |
| DTA | GPT | 0.855 ($\pm 0.018$) | 9.86% | 21.62% |
| | Gemini | 0.917 ($\pm 0.019$) | 51.92% | 10.17% |
| | Haiku | 0.762 ($\pm 0.021$) | 14.4% | 28.96% |
| Intersection | GPT | 0.887 ($\pm 0.016$) | 14.45% | 12.98% |
| | Gemini | 0.887 ($\pm 0.031$) | 45.15% | 19.33% |
| | Haiku | 0.76 ($\pm 0.086$) | 10.71% | 34.09% |

#### Model Diversity

Model diversity plays a central role in ensemble performance [61]. The `Soft-Vote` approach, combining three distinct QA models, benefits from a “healthy” inter-model disagreement (i.e., model heterogeneity [59]), enabling it to balance strengths and mitigate weaknesses across agents. To make ensemble work, it is also important to make good trade-off between maximising model accuracy and maintaining sufficient diversity [62]. `MAD-Soft-Vote` in Table 1 ensembles three debating models, each of which refines its reasoning through peer input before voting. While MAD improves individual model decisions, it reduces overall diversity among different debating models due to convergence in reasoning, demonstrated by the high correlations between `GPT-MAD`, `Gemini-MAD` and `Haiku-MAD`. This leads to marginal gains of `MAD-Soft-Vote` over the best debating models and causes `MAD-Soft-Vote` to underperform `Soft-Vote`. Comparatively, `Adj-Soft-Vote`, which combines two adjudication models, gains significant improvement on Intervention, because of model diversity between the two adjudicators, demonstrated by the fact that the correlation between `Adj-Judge` and `Adj-Rank` on Intervention is much lower than that on DTA (see Figure 2). In summary, while both debating and adjudication offer refinements, preserving model diversity through simple ensembling (`Soft-Vote`) remains the most cost-effective and robust strategy under resource constraints.

### Additional Results: Re-Ranking

Table 4 shows further performance improvement using both macro- and micro-level re-ranking (for simplicity of comparison, *α* = *β* = 0.5). Table 4 also includes two important baselines from [30]. `GPT_Cos_Sim_Criteria` ranks candidate studies based on their semantic relevance with the selection criteria. `GPT_QA_Soft_Both_ReRank` is the best approach in [2] that integrates both macro- and micro-level re-ranking, thus comparable to our re-ranking variants.

**Table 4.**
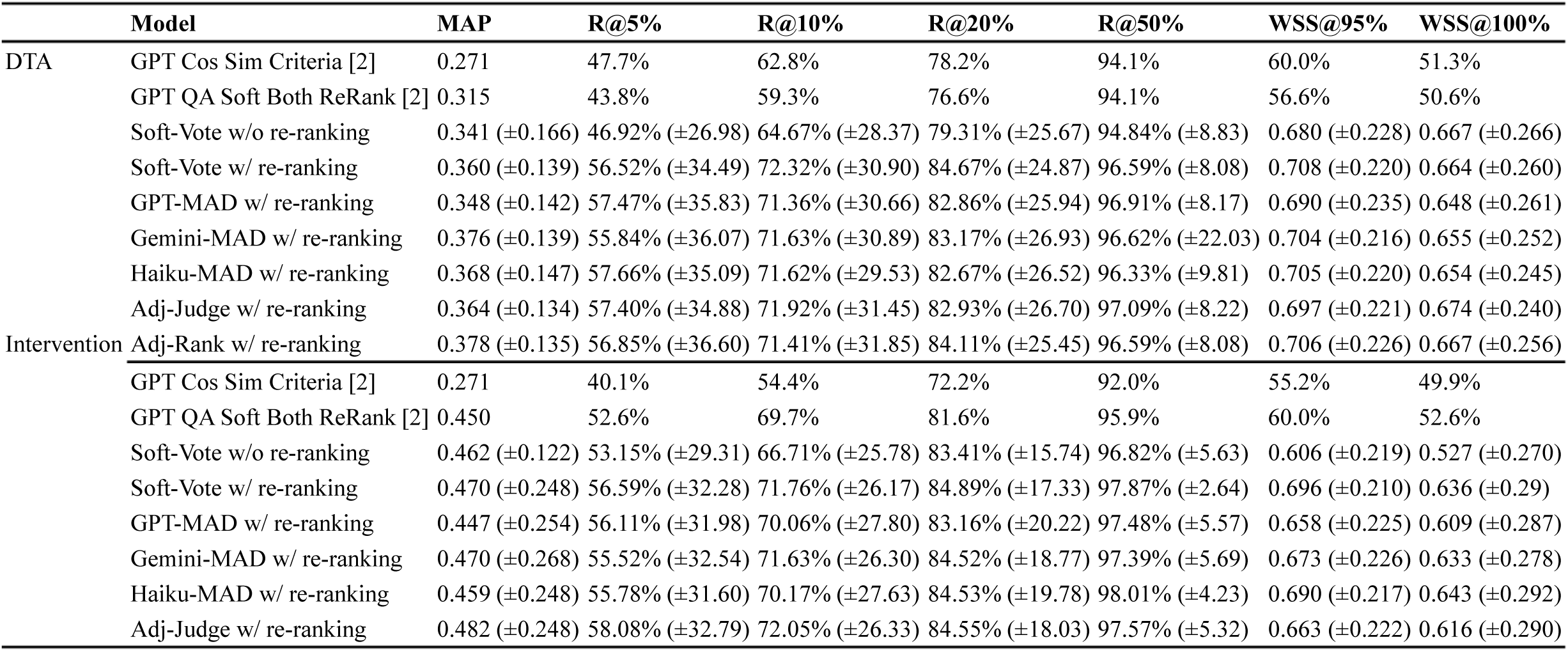
Performance Improvements by Re-ranking.

`Soft-Vote w/o re-ranking` significantly outperformed the best results in [30], particularly in WSS. Although GPT-3.5 was used in [30], we can still claim the benefit of multi-LLM collaboration based on the significant performance gain of `Soft-Vote w/o re-ranking` over the primary models. `GPT_Cos_Sim_Criteria`’s decent performance highlights its plausible “model heterogeneity” that can be leveraged for improving our multi-LLM collaboration approaches through re-ranking. Indeed, `Soft-Vote w/ re-ranking` achieved notable improvements over `Soft-Vote w/o re-ranking` on both DTA and Intervention, reaching new states of the art in term of WSS@95%. Notably, re-ranking also consistently improved the performances of other collaborative strategies by large margins, making them almost rival `Soft-Vote w/ re-ranking` on DTA.

### Limitations

While this study demonstrates the effectiveness of multi-LLM collaboration for abstract screening, several limitations should be acknowledged. Additionally, while our experiments concatenated titles and abstracts for processing, the small subset of title-only records (approximately 2–3% of documents in TAR2019) provided limited context compared to full title-and-abstract screening. This may have slightly affected classification validity for these records, though sensitivity analyses suggested minimal impact on overall performance metrics.

Secondly, our evaluation is confined to the biomedical TAR2019 benchmark, which comprises Cochrane systematic reviews in clinical intervention and diagnostic technology assessment. Medical abstracts typically follow structured formats (e.g., IMRAD: Introduction, Methods, Results, and Discussion) with consistent terminology and well-established reporting standards such as CONSORT (Consolidated Standards of Reporting Trials) for trials and STARD (Standards for Reporting of Diagnostic Accuracy Studies) for diagnostic accuracy studies. These characteristics may make biomedical abstracts particularly amenable to LLM-based classification. The performance of our multi-LLM collaboration strategies in other domains such as social sciences, environmental studies, education research, or humanities where abstracts may be less structured, terminology more varied, and reporting standards less uniform, remains to be validated. Cross-domain evaluation represents an important direction for future work to establish the generalizability of these approaches.

## Conclusion

This study presents an in-depth investigation into LLM-based multi-agent collaborative strategies for automating abstract screening in systematic reviews. We successfully developed and evaluated three collaborative strategies—majority voting, multi-agent debate and LLM-based adjudication, and demonstrated that collaboration among multiple, cost-effective LLMs has high potential to substantially reduce screening workload and cost. The collaborative frameworks effectively mitigate individual LLMs’ biases through collective intelligence.

Majority voting emerged as the most robust solution, achieving consistent and significant performance gains in all settings. Analysis demonstrated the core role of model diversity (i.e., model heterogeneity) on the success of aggregating relatively weaker screening models. While debating improves screening performance, its benefits are more model-specific. Models that stick to their own decisions are less likely benefiting from peers, which is deemed an important insight for developing effective multi-agent debating systems in the context of abstract screening. Nevertheless, we argue that it is worth conducting further research in this strategy with cost-effective lightweight LLMs, especially when taking into consideration its strikingly lower costs than the recent LLM-as-a-Judge paradigm. The economic implications are also substantial. Majority voting only costs less than 1/14 of that of adjudication methods based on a strong LLM, and achieves more than 186× cost reduction compared to single human reviewer based on a conservative estimation according to British academic salary scales. By demonstrating that multi-LLM collaboration can achieve superior performance at a substantially lower cost, this research offers a pathway toward making systematic review automation both more effective and affordable.

## Supporting information

Supplementary File 1 for Implementation Details

## Statement of Data Availability

All data of analysis is published in the main text and the supplementary materials, while all experimental configurations, including model versions and temperature settings, are detailed in the main text. Regarding the raw data for the experiments, the TAR2019 benchmark dataset is available in the CLEF-TAR repository at https://github.com/CLEF-TAR/tar/tree/master/2019-TAR (Task 2 of CLEF eHealth 2019 Technology-Assisted Review). The titles and abstracts for all documents in the TAR2019 dataset are copyrighted but can be extracted from PubMed programmatically via the PubMed API using the provided PubMed IDs (PMIDs) in the CLEF-TAR repository. The selection criteria for each systematic review are included in the CLEF-TAR repository. The converted inclusion criteria questions used in our question-answering framework are provided in Appendix D of Supplementary File 1. To enhance reproducibility, we have created a public GitHub repository containing the prompts used for all collaboration strategies along with implementation details: https://github.com/Ope-Akinseloyin/Multi_LLM-Citation-Screening.

## Data Availability

All data produced in the present study are available upon reasonable request to the authors

## Footnotes

1 Gemini 1.5 Flash does not allow a temperature of 0.0 on certain prompts/abstracts, so 0.1 was selected as the lowest consistent value across all models in our experiments.

