## Supplementary File 1 for Implementation Details for "LLM-based Multi-Agent Collaboration for Abstract Screening towards Automated Systematic Reviews"

### Appendix

The appendix document contains the following sections:

- Sect. A about the prompts for question answering, multi-agent debate and LLM-based adjudication
- Sect. B about an example using one abstract and one inclusion criteria questions.
- Sect. C about further explanation of the difference between the new reasoning and explanation in multi-agent debate.
- Sect. D about more details of the TAR2019 dataset used in our experiments.
- Sect. E about a detailed estimation of human review cost and theoretical financial savings using multi-LLM collaboration.

#### Appendix A: Prompts

Figure A1 below shows the prompt for the QA models.

**Prompt 1: Question Answering**

**Role:** You are a researcher screening titles and abstracts of scientific papers for the systematic review "{review_title}"

**Task:** Analyse the abstract below within the brackets and answer the question below. Taking a step-by-step approach towards reasoning and answering the question. The answer should be in either a positive, neutral, or negative sentiment format.

**Abstract:** {abstract}

**Question:** {question}

**Instruction:** Keep your answers as short as possible. The answer should be in either a positive, neutral, or negative sentiment format. The answer must contain your answer, how you got to your answer (reasoning path), confidence of your answer from 0 to 1 and finally what extra information would make you more confident in your answer. Format of the answer should be like the example below in text(string) not JSON or Code:

(

"Answer": (Positive, Neutral or Negative),

"Reasoning path": str,

"Confidence level": float,

"Extra Information": str,

Figure A1. Prompt for Inclusion Criteria Question Answering.

Figure A2 below shows the prompt for the Multi-Agent Debate models.

#### Prompt 2: Multi-Agent Debate

**Role:** You are a researcher screening titles and abstracts of scientific papers for the systematic review "{review_title}"

**Task:** Analyse the abstract below within the brackets and answer the question below. Taking a step-by-step approach towards reasoning and answering the question.
Also take into consideration your previous answer, confidence and reason. As well as the answers, confidence and reasons giving to you by your colleagues Gemini and Claude.

**Question:** {question}

**Abstract:** {abstract}

**Your Initial Answer:** {initial_answer}

**Claude Answer:** {claude_answer}

**Gemini Answer:** {gemini_answer}

**Instruction:** Keep your answers as short as possible. The answer should be in either a positive, neutral, or negative sentiment format. The answer must contain your answer, how you got to your answer (reasoning path), confidence of your answer from 0 to 1 and finally what extra information would make you more confident in your answer. Format of the answer should be like the example below in text(string) not JSON or Code:

(

"Answer": (Positive, Neutral or Negative),

"Reasoning path": str,

"Confidence level": float,

"Extra Information": str,

"Does your Previous Answer Change": (Yes, No),

Figure A2. Prompt for Multi-Agent Debate for Answering Refinement.

Figure A3 below shows the prompt for the LLM-based Adjudication models.

#### Prompt 3: LLM-based Adjudication

**Role:** You are a Senior researcher in a group screening titles and abstracts of scientific papers for the systematic review '{review_title}'

**Task:** Analyse the abstract below within the brackets and choose the best answer produced by the other researchers in your group for the question below. Taking a step-by-step approach towards reasoning and answering the question.

Also take into answers, confidence and reason by your junior colleagues Gemini, GPT and Claude. Avoid any position biases and ensure that the order in which the responses were presented does not influence your decision.

Do not allow the length of the responses to influence your evaluation. Do not favor certain names of the assistants. Be as objective as possible.

**Gemini's Answer:** {gemini_answer}

**Claude's Answer:** {claude_answer}

**GPT's Answer:** {gpt_answer}

**Question:** {question}

**Abstract:** {abstract}

**Instruction:** Keep your answers as short as possible. The answer should be in either a positive, neutral, or negative sentiment format. The answer should contain your answer, how you got to your answer (reasoning path), confidence of your answer from 0 to 100 and finally what extra information would make you more confident in your answer. Please rate the response on a scale of 0 to 1. Format of the answer must be like the example below in text(string) not JSON or Code:

(

"Answer": (Positive, Neutral or Negative),

"Reasoning path": str,

"Confidence level": float,

"Extra Information": str,

"Gemini's Rating": float,

"Claude's Rating": float,

"GPT's Rating": float,

"Which of your colleagues has the best answer": str,

"Reason for best answer": str,

"Which of your colleagues has the worst answer": str,

Figure A3. Prompt for LLM-based Adjudication.

#### Appendix B: Example of Multi-Agent Collaboration Strategies for Screening Prioritization

Figure A4 shows an example, including the title and abstract of the abstract of a candidate study, and an inclusion criterion (converted to an inclusion criteria question).

**Question:**

1. Does the study involve full-text diagnostic test accuracy research on any individual index test or a combination of tests against a reference standard?

**Title:**

Neck circumference to thyromental distance ratio: a new predictor of difficult intubation in obese patients.

**Abstract:**

Background: This study was performed to assess whether intubation is more difficult in obese patients and to assess the ability of a new index: the ratio of the neck circumference to thyromental distance (NC/TM), to predict difficult intubation in obese patients.

Methods: The incidence of difficult tracheal intubation in 123 obese (BMI≥27.5 kg m(-2)) and 125 non-obese patients was compared. Difficult intubation was determined using the intubation difficulty scale (IDS≥5). The NC/TM ratio was calculated and its ability to predict difficult intubation in obese patients was compared with that of established predictors including high BMI, the Mallampati score, the Wilson score, NC, width of mouth opening, sternomental distance, TM, and a previous history of difficult intubation.

Results: Difficult intubation was more frequent in obese patients than in non-obese patients (13.8% vs 4.8%; P=0.016). Multivariate analysis revealed that the Mallampati score, the Wilson score, and NC/TM independently predicted difficult intubation in obese patients. Among these three indices, NC/TM showed the highest sensitivity and a negative predictive value, and largest area under the curve on an ROC curve.

Conclusions: Difficult intubation was more common in obese patients and the NC/TM was a better method for predicting difficult intubation than other established indices.

Figure A4. An Example to Illustrate Question Answering-based Abstract Screening.

Figure A5 shows the output of the question answering model.

Output 1: Question Answering

(

"Answer": "Negative",

"Reasoning path": "The abstract describes a retrospective study assessing the prevalence of vascular disease in patients with bronchiectasis, without any mention of inhaled corticosteroids or a randomized controlled trial design. It focuses on vascular disease risk factors rather than treatment comparisons. ",

"Confidence level": 95,

"Extra Information": "Access to the full text of the study to confirm the absence of any treatment-related comparisons would increase confidence."

)

Figure A5. An Example Output of the Question Answering Model.

Figure A6 shows the output of the multi-agent debate model.

Output 2: Multi-Agent Debate

(

"Answer": "Negative",

"Reasoning path": "The abstract clearly indicates that the study is a retrospective observational study focused on vascular disease in bronchiectasis patients, with no mention of inhaled corticosteroids or a randomized controlled trial design. This aligns with the assessments made by colleagues.",

"Confidence level": 95,

"Extra Information": "Access to the full text of the study to confirm the absence of any treatment-related comparisons would increase confidence.'

"Does your Previous Answer Change": "No",

"Why for the change or not in your previous answer": The reasoning and conclusion remain consistent with the previous analysis and those of colleagues, confirming the absence of an RCT involving ICS."

)

Figure A6. An Example Output of the Multi-Agent Debate Model.

Figure A7 shows the output of the LLM-based adjudication model.

**Output 3: LLM-based Adjudication**

(

"Answer": "Negative",

"Reasoning path": "The abstract explicitly describes a retrospective study examining the prevalence and risk factors of vascular disease in bronchiectasis patients. There is no mention of any intervention, let alone an RCT involving ICS."

"Confidence level": 1.0,

"Extra Information": "None needed. The abstract is clear about the study design and purpose."

"Gemini's Rating": 1.0,

"Claude's Rating": 0.9,

"GPT's Rating": 0.9,

"Which of your colleagues has the best answer": "Gemini",

"Reason for best answer": "Gemini correctly identifies the retrospective nature as the key reason why this cannot be an RCT, and does so concisely and with high confidence."

"Which of your colleagues has the worst answer": "Claude",

"Reason for worst answer": "While Claude's answer is correct, suggesting a need to review the full text is unnecessary. The abstract provides sufficient information to definitively rule out an RCT. GPT's answer is similar to Claude's but slightly better as it acknowledges the focus on risk factors rather than treatment."

Figure A7. An Example Output of the LLM-based Adjudication Model.

#### Appendix C: Difference between Reasoning and Explanation in Multi-Agent Debate

In Figure A6, the debating agent not only gives a (new) reasoning path, but also an explanation for "Why for the change or not in your previous answer". Further explanations are given below.

New Reasoning: This is the full reasoning path the debating model provides after reconsidering its answer in light of its peers' input. It can be a general justification drawn from the abstract, possibly incorporating peers' perspectives, but it does not have to explain why the answer changed (or didn't change) — it's simply the updated rationale for the current answer.

Explanation for Answer Change: This is specifically the model's explicit explanation of why its current answer differs from its original answer, focusing on what new information or peer input prompted the change (or confirmation).

If the answer changed: it states what prompted the change (e.g., a new detail from the abstract, a peer's argument).

If the answer didn't change: it explains why the original answer still holds despite reviewing peers' perspectives (e.g., no new evidence strong enough to overturn the initial decision).

Below gives an example for Answer change:

(

"Answer": "Positive",

"Reasoning path": "The abstract describes a study that evaluates the diagnostic accuracy of multiple airway physical examination tests against the reference standard of difficult tracheal intubation. It provides sensitivity and specificity values for these tests, indicating a thorough assessment of their accuracy, which aligns with the systematic review's objective. "

"Confidence level": 0.90,

"Extra Information": "Access to the full text to confirm the reference standard and methodology would increase confidence."

"Does your Previous Answer Change": "Yes",

"Why for the change or not in your previous answer": The additional insights from colleagues Claude and Gemini highlight the study's alignment with the systematic review's criteria, which I initially overlooked regarding the reference standard and diagnostic accuracy assessment."

#### Appendix D: Details of the TAR2019 Dataset

Table A1: Information of each SR within the DTA Category

| Topic ID | Title | # total PMIDs | # abs rel | % abs rel |
|---|---|---|---|---|
| CD012567 | Positron emission tomography (PET) and magnetic resonance imaging (MRI) for assessing tumour resectability in advanced epithelial ovarian/fallopian tube/primary peritoneal cancer | 5863 | 11 | 0.002 |
| CD012669 | Point-of-care ultrasonography for diagnosing thoracoabdominal injuries in patients with blunt trauma | 1153 | 70 | 0.061 |
| CD012233 | Transabdominal ultrasound and endoscopic ultrasound for diagnosis of gallbladder polyps | 430 | 38 | 0.088 |
| CD008874 | Airway physical examination tests for detection of difficult airway management in apparently normal adult patients | 1799 | 114 | 0.063 |
| CD012768 | Xpert MTB/RIF assay for extrapulmonary tuberculosis and rifampicin resistance | 119 | 43 | 0.361 |
| CD012080 | Non-invasive diagnostic tests for Helicobacter pylori infection | 6314 | 73 | 0.012 |
| CD011686 | Triage tools for detecting cervical spine injury in pediatric trauma patients | 8032 | 63 | 0.008 |
| CD009044 | Diagnostic tests for autism spectrum disorder (ASD) in preschool children | 3120 | 11 | 0.004 |

Table A2: Information of each SR within the Intervention Category

| Topic ID | Title | # total PMIDs | # abs rel | % abs rel |
|---|---|---|---|---|
| CD010239 | Lower versus higher oxygen concentrations titrated to target oxygen saturations during resuscitation of preterm infants at birth | 221 | 12 | 0.054 |
| CD012551 | Non-pharmacological interventions for treating chronic prostatitis/chronic pelvic pain syndrome | 555 | 66 | 0.119 |
| CD011571 | Antistreptococcal interventions for guttate and chronic plaque psoriasis | 128 | 11 | 0.086 |
| CD011140 | Implantable miniature telescope (IMT) for vision loss due to end-stage age-related macular degeneration | 280 | 4 | 0.014 |
| CD012455 | Melatonin for the promotion of sleep in adults in the intensive care unit | 1570 | 7 | 0.004 |
| CD009642 | Continuous intravenous perioperative lidocaine infusion for postoperative pain and recovery in adults | 1869 | 62 | 0.033 |
| CD007867 | Prescribed hypocaloric nutrition support for critically-ill adults | 918 | 17 | 0.019 |
| CD011768 | Educational interventions for improving primary caregiver complementary feeding practices for children aged 24 months and under | 8963 | 53 | 0.006 |
| CD011977 | Blue-light filtering intraocular lenses (IOLs) for protecting macular health | 192 | 49 | 0.255 |
| CD012164 | Subfascial endoscopic perforator surgery (SEPS) for treating venous leg ulcers | 60 | 7 | 0.117 |
| CD010038 | Face-to-face interventions for informing or educating parents about early childhood vaccination | 8616 | 23 | 0.003 |
| CD009069 | Prophylactic vaccination against human papillomaviruses to prevent cervical cancer and its precursors | 1664 | 78 | 0.047 |
| CD001261 | Vaccines for preventing typhoid fever | 497 | 67 | 0.135 |
| CD010753 | Antidepressants for insomnia in adults | 2247 | 28 | 0.012 |
| CD006468 | Anticoagulation for people with cancer and central venous catheters | 3483 | 50 | 0.014 |
| CD010558 | Psychological therapies for treatment-resistant depression in adults | 2770 | 37 | 0.013 |
| CD000996 | Inhaled corticosteroids for bronchiectasis | 281 | 9 | 0.032 |
| CD012069 | Methylphenidate for attention deficit hyperactivity disorder (ADHD) in children and adolescents âĂŞ assessment of adverse events in non-randomised studies | 2903 | 283 | 0.097 |
| CD004414 | Interventions for preventing occupational irritant hand dermatitis | 336 | 16 | 0.048 |
| CD012342 | Comparison of a therapeutic-only versus prophylactic platelet transfusion policy for people with congenital or acquired bone marrow failure disorders | 2294 | 6 | 0.003 |

Table A3: Selection criteria and Questions generated for the SRs in the DTA Category

| Topic | Selection Criteria | Questions |
|---|---|---|
| **CD012567** | Diagnostic accuracy studies addressing the accuracy of preoperative FDG-PET/CT, conventional or DW-MRI on assessing tumour resectability in women with advanced stage (III to IV) epithelial ovarian/fallopian tube/primary peritoneal cancer who are scheduled to undergo primary debulking surgery. | Does the study evaluate the diagnostic accuracy of preoperative FDG-PET/CT in assessing tumour resectability in women with advanced stage (III to IV) epithelial ovarian/fallopian tube/primary peritoneal cancer? |
| | | Does the study evaluate the diagnostic accuracy of conventional MRI in assessing tumour resectability in women with advanced stage (III to IV) epithelial ovarian/fallopian tube/primary peritoneal cancer? |
| | | Does the study evaluate the diagnostic accuracy of DW-MRI in assessing tumour resectability in women with advanced stage (III to IV) epithelial ovarian/fallopian tube/primary peritoneal cancer? |
| | | Does the study focus on women who are scheduled to undergo primary debulking surgery? |
| | | Does the study specifically address the accuracy of the mentioned imaging techniques (FDG-PET/CT, conventional or DW-MRI) in preoperative assessment of tumour resectability in women with advanced stage (III to IV) epithelial ovarian/fallopian tube/primary peritoneal cancer? |
| **CD012669** | We assessed studies for eligibility using predefined inclusion and exclusion criteria. We included either prospective or retrospective diagnostic cohort studies that enrolled patients of any age and gender who sustained any type of blunt injury in a civilian scenario. Eligible studies had to provide sufficient information to construct a 2 x 2 table of diagnostic accuracy to allow for calculating sensitivity, specificity, and other indices of diagnostic test accuracy. | Does the study involve a diagnostic cohort study? |
| | | Did the study enroll patients who sustained blunt injuries? |
| | | Does the study include patients of any age and gender? |
| | | Is the study conducted in a civilian scenario? |
| | | Does the study provide sufficient information to construct a 2 x 2 table of diagnostic accuracy? |
| **CD012233** | Studies reporting on the diagnostic accuracy data (true positive, false positive, false negative and true negative) of the index test (TAUS or EUS or both) for detection of gallbladder polyps, differentiation between true and pseudo polyps, or differentiation between dysplastic polyps/carcinomas and adenomas/pseudo polyps. We only accepted histopathology after cholecystectomy as the reference standard, except for studies on | Does the study report on the diagnostic accuracy of TAUS or EUS or both for the detection of gallbladder polyps? |
| | | Does the study report on the diagnostic accuracy of TAUS or EUS or both for the differentiation between true and pseudo polyps? |
| | | Does the study report on the diagnostic accuracy of TAUS or EUS or both for the differentiation between dysplastic polyps/carcinomas and adenomas/pseudo polyps? |
| | | Does the study use histopathology after cholecystectomy as the reference standard for diagnostic accuracy? |

| | | |
|---|---|---|
| | diagnosis of gallbladder polyp. For the latter studies, we also accepted repeated imaging up to six months by TAUS or EUS as the reference standard. | For studies on the diagnosis of gallbladder polyps, does the study also accept repeated imaging up to six months by TAUS or EUS as the reference standard? |
| **CD008874** | We considered full-text diagnostic test accuracy studies of any individual index test, or a combination of tests, against a reference standard. Participants were adults without obvious airway abnormalities, who were having laryngoscopy performed with a standard laryngoscope and the trachea intubated with a standard tracheal tube. Index tests included the Mallampati test, modified Mallampati test, Wilson risk score, thyromental distance, sternomental distance, mouth opening test, upper lip bite test, or any combination of these. The target condition was difficult airway, with one of the following reference standards: difficult face mask ventilation, difficult laryngoscopy, difficult tracheal intubation, and failed intubation. | Does the paper investigate the diagnostic accuracy of any individual index test or combination of tests against a reference standard? |
| | | Are the participants in the study adults without obvious airway abnormalities? |
| | | Were laryngoscopy and tracheal intubation performed using standard equipment (laryngoscope and tracheal tube)? |
| | | Does the paper include the evaluation of any of the following index tests: Mallampati test, modified Mallampati test, Wilson risk score, thyromental distance, sternomental distance, mouth opening test, upper lip bite test, or any combination of these? |
| | | Does the paper focus on the target condition of difficult airway, with one of the following reference standards: difficult face mask ventilation, difficult laryngoscopy, difficult tracheal intubation, or failed intubation? |
| **CD012768** | We included diagnostic accuracy studies of Xpert in people presumed to have extrapulmonary TB. We included TB meningitis and pleural, lymph node, bone or joint, genitourinary, peritoneal, pericardial, and disseminated TB. We used culture as the reference standard. For pleural TB, we also included a composite reference standard, which defined a positive result as the presence of granulomatous inflammation or a positive culture result. For rifampicin resistance, we used culture-based drug susceptibility testing or MTBDRplus as the reference standard. | Does the study focus on the diagnostic accuracy of Xpert in people with extrapulmonary TB? |
| | | Does the study include patients with TB meningitis, pleural, lymph node, bone or joint, genitourinary, peritoneal, pericardial, or disseminated TB? |
| | | Does the study use culture as the reference standard for diagnosing extrapulmonary TB? |
| | | Does the study include a composite reference standard for diagnosing pleural TB, which considers the presence of granulomatous inflammation or a positive culture result as a positive result? |
| | | Does the study use culture-based drug susceptibility testing or MTBDRplus as the reference standard for rifampicin resistance? |
| **CD012080** | We included diagnostic accuracy studies that evaluated at least one of the index tests (urea breath test using isotopes such as 13C or 14C, | Does the study evaluate the diagnostic accuracy of at least one of the index tests (urea breath test using isotopes such as 13C or 14C, serology, or stool antigen test |

| | | |
|---|---|---|
| | serology and stool antigen test) against the reference standard (histopathological examination using H & E stain, special stains or immunohistochemical stain) in people suspected of having H pylori infection. | Is the reference standard used in the study histopathological examination using H & E stain, special stains, or immunohistochemical stain? |
| | | Are the participants in the study suspected of having H pylori infection? |
| | | Does the study include human participants? |
| | | Is the study design focused on diagnostic accuracy evaluation rather than other aspects of H pylori infection? |
| **CD011686** | We included all retrospective and prospective studies involving children following blunt trauma that evaluated the accuracy of the NEXUS criteria, the Canadian C-spine Rule, or both. Plain radiography, computed tomography (CT) or magnetic resonance imaging (MRI) of the cervical spine, and follow-up were considered as adequate reference standards. | Does the study involve children who have experienced blunt trauma? |
| | | Does the study evaluate the accuracy of the NEXUS criteria? |
| | | Does the study evaluate the accuracy of the Canadian C-spine Rule? |
| | | Does the study evaluate the accuracy of both the NEXUS criteria and the Canadian C-spine Rule? |
| | | Does the study use plain radiography, computed tomography (CT), or magnetic resonance imaging (MRI) of the cervical spine as reference standards for evaluation? |
| **CD009044** | Publications had to: 1. report diagnostic test accuracy for any of the following six included diagnostic tools: Autism Diagnostic Interview - Revised (ADI-R), Gilliam Autism Rating Scale (GARS), Diagnostic Interview for Social and Communication Disorder (DISCO), Developmental, Dimensional, and Diagnostic Interview (3di), Autism Diagnostic Observation Schedule - Generic (ADOS), and Childhood Autism Rating Scale (CARS); 2. include children of preschool age (under six years of age) suspected of having an ASD; and 3. have a multi-disciplinary assessment, or similar, as the reference standard.Eligible studies included cohort, cross-sectional, randomised test accuracy, and case-control studies. The target condition was ASD. | Does the paper report diagnostic test accuracy for any of the six included diagnostic tools: ADI-R, GARS, DISCO, 3di, ADOS, and CARS? |
| | | Are the participants in the study children of preschool age (under six years of age) suspected of having an ASD? |
| | | Does the paper describe a multi-disciplinary assessment or similar as the reference standard for diagnosing ASD? |
| | | Is the study design of the paper a cohort, cross-sectional, randomized test accuracy, or case-control study? |
| | | Does the paper focus on the target condition of ASD? |

Table A4: Selection criteria and Questions generated for the SRs in the Intervention Category

| Topic | Selection Criteria | Questions |
|---|---|---|
| **CD000996** | All randomised controlled trials (RCTs) comparing ICS with a placebo or no medication. We included children and adults with clinical or radiographic evidence of bronchiectasis, but excluded people with cystic fibrosis. | Is the study design a randomized controlled trial (RCT)? |
| | | Does the study compare the use of inhaled corticosteroids (ICS) with a placebo or no medication? |
| | | Does the study include participants with clinical or radiographic evidence of bronchiectasis? |
| | | Does the study include both children and adults? |
| | | Does the study exclude individuals with cystic fibrosis? |
| **CD001261** | Randomized and quasi-randomized controlled trials (RCTs) comparing typhoid fever vaccines with other typhoid fever vaccines or with an inactive agent (placebo or vaccine for a different disease) in adults and children. Human challenge studies were not eligible. | Is the study design a randomized or quasi-randomized controlled trial? |
| | | Does the study compare typhoid fever vaccines with other typhoid fever vaccines? |
| | | Does the study compare typhoid fever vaccines with an inactive agent (placebo or vaccine for a different disease)? |
| | | Does the study include participants who are adults and children? |
| | | Was the study conducted on human subjects as part of a challenge study? |
| **CD004414** | We included parallel and cross-over randomised controlled trials (RCTs) which examined the effectiveness of barrier creams, moisturisers, gloves, or educational interventions compared to no intervention for the primary prevention of OIHD under field conditions. | Is the study design a randomised controlled trial (RCT)? |
| | | Does the study compare the effectiveness of barrier creams, moisturisers, gloves, or educational interventions? |
| | | Is the study focused on the primary prevention of occupational irritant hand dermatitis (OIHD)? |
| | | Are the interventions being compared to a no intervention control group? |
| | | Were the interventions tested under field conditions? |
| **CD006468** | Randomized controlled trials (RCTs) assessing the benefits and harms of unfractionated heparin (UFH), low-molecular-weight heparin (LMWH), vitamin K antagonists (VKA), or fondaparinux or comparing the effects of two of these anticoagulants in people with cancer and a CVC. | Is the study design a randomized controlled trial (RCT)? |
| | | Does the study assess the benefits and harms of unfractionated heparin (UFH), low-molecular-weight heparin (LMWH), vitamin K antagonists (VKA), or fondaparinux? |
| | | Does the study include participants with cancer? |
| | | Does the study involve people with a central venous catheter (CVC)? |
| | | Does the study compare the effects of two of the mentioned anticoagulants? |
| **CD007867** | We included randomized and quasi-randomized controlled trials comparing hypocaloric nutrition support to normo- or hypercaloric nutrition support or no nutrition support (e.g. fasting) in adults hospitalized in intensive care units (ICUs). | Is the study design a randomized or quasi-randomized controlled trial? |
| | | Does the study compare hypocaloric nutrition support to normo- or hypercaloric nutrition support or no nutrition support? |
| | | Are the participants of the study adults? |
| | | Were the participants hospitalized in intensive care units (ICUs)? |
| | | Does the study focus on the effects of nutrition support on hospitalized adults in ICUs? |

| | | |
|---|---|---|
| **CD009069** | Randomised controlled trials comparing efficacy and safety in females offered HPV vaccines with placebo (vaccine adjuvants or another control vaccine). | Is the study design a randomized controlled trial? |
| | | Does the study compare the efficacy and safety of HPV vaccines? |
| | | Are the participants exclusively females? |
| | | Is the control group receiving either a placebo, vaccine adjuvants, or another control vaccine? |
| | | Does the study report on the comparison of efficacy and safety outcomes between the intervention and control groups? |
| **CD009642** | We included randomized controlled trials comparing the effect of continuous perioperative IV lidocaine infusion either with placebo, or no treatment, or with thoracic epidural analgesia (TEA) in adults undergoing elective or urgent surgery under general anaesthesia. The IV lidocaine infusion must have been started intraoperatively, prior to incision, and continued at least until the end of surgery. | Does the study involve a randomized controlled trial design? |
| | | Does the study compare the effect of continuous perioperative IV lidocaine infusion? |
| | | Does the study compare IV lidocaine infusion with placebo or no treatment? |
| | | Does the study compare IV lidocaine infusion with thoracic epidural analgesia (TEA)? |
| | | Was the IV lidocaine infusion started intraoperatively, prior to incision, and continued until the end of surgery? |
| **CD010038** | We included randomised controlled trials (RCTs) and cluster-RCTs evaluating the effects of face-to-face interventions delivered to parents or expectant parents to inform or educate them about early childhood vaccination, compared with control or with another face-to-face intervention. The World Health Organization recommends that children receive all early childhood vaccines, with the exception of human papillomavirus vaccine (HPV), which is delivered to adolescents. | Is the study design a randomized controlled trial (RCT) or a cluster-RCT? |
| | | Does the study evaluate the effects of face-to-face interventions? |
| | | Are the interventions delivered to parents or expectant parents? |
| | | Do the interventions aim to inform or educate parents about early childhood vaccination? |
| | | Is the study comparing the interventions with either a control group or another face-to-face intervention? |
| **CD010239** | We included randomised controlled trials (including cluster- and quasi-randomised trials) which enrolled preterm infants requiring resuscitation following birth and allocated them to receive either lower ($FiO2 < 0.4$) or higher ($FiO2 \geq 0.4$) initial oxygen concentrations titrated to target oxygen saturation. | Was the study a randomised controlled trial? |
| | | Did the study enroll preterm infants requiring resuscitation following birth? |
| | | Were the infants allocated to receive either lower ($FiO2 < 0.4$) or higher ($FiO2 \geq 0.4$) initial oxygen concentrations? |
| | | Was the oxygen concentration titrated to target oxygen saturation? |
| | | Did the study include cluster- or quasi-randomised trials? |
| **CD010558** | We included randomised controlled trials (RCTs) with | Does the study involve participants aged 18 to 74 years diagnosed with unipolar depression? |

| | | |
|---|---|---|
| | participants aged 18 to 74 years diagnosed with unipolar depression that had not responded to minimum four weeks of antidepressant treatment at a recommended dose. We excluded studies of drug intolerance. Acceptable diagnoses of unipolar depression were based onthe Diagnostic and Statistical Manual of Mental Disorders (DSM-IV-TR) or earlier versions, International Classification of Diseases (ICD)-10, Feighner criteria, or Research Diagnostic Criteria. We included the following comparisons.1. Any psychological therapy versus antidepressant treatment alone, or another psychological therapy.2. Any psychological therapy given in addition to antidepressant medication versus antidepressant treatment alone, or a psychological therapy alone.Primary outcomes required were change in depressive symptoms and number of dropouts from study or treatment (as a measure of acceptability). | Did the participants not respond to a minimum of four weeks of antidepressant treatment at a recommended dose? |
| | | Is the study a randomised controlled trial (RCT)? |
| | | Does the study compare any psychological therapy versus antidepressant treatment alone or another psychological therapy? |
| | | Does the study report primary outcomes related to change in depressive symptoms and number of dropouts from study or treatment? |
| **CD010753** | Randomised controlled trials (RCTs) of adults (aged 18 years or older) with a primary diagnosis of insomnia and all participant types including people with comorbidities. Any antidepressant as monotherapy at any dose whether compared with placebo, other medications for insomnia (e.g. benzodiazepines and 'Z' drugs), a different antidepressant, waiting list control or treatment as usual. | Is the study design a randomized controlled trial (RCT)? |
| | | Does the study involve adults aged 18 years or older? |
| | | Do the participants have a primary diagnosis of insomnia? |
| | | Does the study include participants with comorbidities? |
| | | Is the intervention being studied an antidepressant used as monotherapy at any dose? |
| **CD011140** | We planned to include randomized controlled trials (RCTs) and quasi-randomized trials that compared the IMT versus no IMT. | Is the study design a randomized controlled trial (RCT) or quasi-randomized trial? |
| | | Does the study compare the use of inspiratory muscle training (IMT) with no IMT? |
| | | Is the intervention in the study focused on training the inspiratory muscles? |
| | | Does the study include a control group that does not receive IMT? |

| | | |
|---|---|---|
| | | Are the outcomes of interest compared between the IMT group and the no IMT group? |
| **CD011571** | Randomised trials of one or more antistreptococcal interventions in patients with guttate or chronic plaque psoriasis. | Does the study involve a randomised trial design? |
| | | Does the study focus on patients with guttate or chronic plaque psoriasis? |
| | | Does the study investigate one or more antistreptococcal interventions? |
| | | Is the study conducted on human subjects? |
| | | Does the study report outcomes related to the effectiveness or efficacy of the antistreptococcal interventions in patients with guttate or chronic plaque psoriasis? |
| **CD011768** | Randomised controlled trials (RCTs), comparing educational interventions to no intervention, usual practice, or educational interventions provided in conjunction with another intervention, so long as the educational intervention was only available in the experimental group and the adjunctive intervention was available to the control group. Study participants included caregivers of infants aged 4 to 24 months undergoing complementary feeding. Pregnant women who were expected to give birth and commence complementary feeding during the period of the study were also included. | Is the study design a randomized controlled trial (RCT)? |
| | | Does the study compare educational interventions to either no intervention or usual practice? |
| | | Was the educational intervention only available in the experimental group? |
| | | Was the adjunctive intervention available to the control group? |
| | | Were the study participants caregivers of infants aged 4 to 24 months undergoing complementary feeding, or pregnant women expected to give birth and commence complementary feeding during the study period? |
| **CD011977** | We included randomised controlled trials (RCTs), involving adult participants undergoing cataract extraction, where a blue-light filtering IOL was compared with an equivalent non-blue-light filtering IOL. | Is the study design a randomized controlled trial (RCT)? |
| | | Does the study involve adult participants? |
| | | Does the study focus on cataract extraction? |
| | | Is the comparison between a blue-light filtering IOL and a non-blue-light filtering IOL? |
| | | Is the study evaluating the effects of the blue-light filtering IOL on the outcomes? |
| **CD012069** | We included non-randomised study designs. These comprised comparative and non-comparative cohort studies, patient-control studies, patient reports/series and cross-sectional studies of methylphenidate administered at any dosage or formulation. We also included methylphenidate groups from RCTs assessing methylphenidate versus other | Does the study design include non-randomized comparative and non-comparative cohort studies, patient-control studies, patient reports/series, and cross-sectional studies? |
| | | Does the study involve the administration of methylphenidate at any dosage or formulation? |
| | | Does the study include methylphenidate groups from randomized controlled trials (RCTs) comparing methylphenidate with other interventions for ADHD? |
| | | Does the study include data from follow-up periods in RCTs? |

| | | |
|---|---|---|
| | interventions for ADHD as well as data from follow-up periods in RCTs. Participants had to have an ADHD diagnosis (from the 3rd to the 5th edition of the Diagnostic and Statistical Manual of Mental Disorders or the 9th or 10th edition of theInternational Classification of Diseases, with or without comorbid diagnoses. We required that at least 75% of participants had a normal intellectual capacity (intelligence quotient of more than 70 points) and were aged below 20 years. We excluded studies that used another ADHD drug as a co-intervention. | Do the participants in the study have a diagnosis of ADHD according to the 3rd to the 5th edition of the Diagnostic and Statistical Manual of Mental Disorders or the 9th or 10th edition of the International Classification of Diseases, with or without comorbid diagnoses? |
| **CD012164** | We included randomised controlled trials (RCTs) of interventions that examined the use of SEPS independently or in combination with another intervention for the treatment of venous leg ulcers. | Is the study design a randomized controlled trial (RCT)? |
| | | Does the study focus on interventions for the treatment of venous leg ulcers? |
| | | Does the study examine the use of SEPS (Sequential Elastic Pressure System) as an intervention? |
| | | Does the study examine the use of SEPS in combination with another intervention? |
| | | Is the study independent and not a duplicate of any other included study? |
| **CD012342** | We included RCTs, non-RCTs, and CBAs that involved the transfusion of platelet concentrates (prepared either from individual units of whole blood or by apheresis any dose, frequency, or transfusion trigger) and given to treat or prevent bleeding among people with congenital or acquired bone marrow failure disorders. | Does the study involve the transfusion of platelet concentrates prepared from individual units of whole blood or by apheresis? |
| | | Is the study focused on treating or preventing bleeding among people with congenital or acquired bone marrow failure disorders? |
| | | Does the study include randomized controlled trials (RCTs)? |
| | | Does the study include non-randomized controlled trials (non-RCTs)? |
| | | Does the study include controlled before-and-after studies (CBAs)? |
| **CD012455** | We included randomized and quasi-randomized controlled trials with adult participants (over the age of 16) admitted to the ICU with any diagnoses given melatonin versus a comparator to promote overnight sleep. We included participants who were mechanically ventilated and those who were not mechanically ventilated. We planned to include studies that compared the use of melatonin, given at an appropriate | Is the study a randomized or quasi-randomized controlled trial? |
| | | Does the study involve adult participants (over the age of 16) admitted to the ICU? |
| | | Does the study compare the use of melatonin with a comparator to promote overnight sleep? |
| | | Does the study include participants who are mechanically ventilated? |
| | | Does the study compare the use of melatonin against no agent or against another agent specifically administered to promote sleep? |

| | | |
|---|---|---|
| | clinical dose with the intention of promoting night-time sleep, against no agent; or against another agent administered specifically to promote sleep. | |
| **CD012551** | We included randomized controlled trials in men with a diagnosis of CP/CPPS. We included all available non-pharmacological interventions. Two review authors independently classified studies and abstracted data from the included studies, performed statistical analyses and rated quality of evidence (QoE) according to the Grading of Recommendations Assessment, Development and Evaluation methods. The primary outcomes were prostatitis symptoms and adverse events. The secondary outcomes were sexual dysfunction, urinary symptoms, quality of life, anxiety and depression. | Is the study a randomized controlled trial?<br>Does the study involve men diagnosed with chronic prostatitis/chronic pelvic pain syndrome (CP/CPPS)?<br>Does the study investigate non-pharmacological interventions?<br>Were the studies independently classified and data abstracted by two review authors?<br>Does the study report outcomes related to prostatitis sym or adverse events? |

#### Appendix E: Detailed Breakdown of Estimation of Costs for Human Reviewer and Theoretical Financial Savings Using Multi-LLM Collaboration

All three strategies will bring substantial cost-down compared to the expensive human screening process. Based on the most conservative assumptions that a postgraduate research assistant-level reviewer screens 30-60 abstracts per hour (at a reading rate of 1—2 per minutes according to the most up-to-date Cochrane Handbook [1], which is actually a reading rate only achievable by experienced domain experts) and the minimal hourly rate is £14, then it will cost approximately 1,110—2,220 hours and £15540—£31080 to screen all 666,22 abstracts, which are about $20,639—$41277 according to the exchange rate on 3 Aug 2025. Compared with Table A5, even the most expensive Adjudicator approach represents only about 3.9% to 7.8% of the lower-bound human screening cost, highlighting the significant economic viability of LLM-based screening automation. Foreseeing stronger performances and lower costs of using even commercial LLMs, the economical gain is expected to become bigger in the near future.

Table A5: LLM Pricing and Cost Breakdown (copied from Table 2 in the main text)

| Setting | Type | Model | Input Price | Input Cost | Output Price | Output Cost | Total Cost |
|---|---|---|---|---|---|---|---|
| DTA | Question Answering | GPT-4o Mini | 0.15 | $2.54 | 0.6 | $8.54 | $11.08 |
| | | Gemini 1.5 Flash | 0.075 | $1.27 | 0.3 | $4.17 | $5.44 |
| | | Claude 3 Haiku | 0.25 | $4.24 | 1.25 | $23.66 | $27.90 |
| | Majority Voting | Soft-Vote | - | $8.05 | - | $36.38 | $44.43 |
| | Debating | GPT-4o Mini | 0.15 | $18.32 | 0.6 | $47.38 | $65.70 |
| | | Gemini 1.5 Flash | 0.075 | $13.19 | 0.3 | $42.64 | $55.82 |
| | | Claude 3 Haiku | 0.25 | $25.16 | 1.25 | $65.15 | $90.31 |
| | Voting on debating | MAD-Soft-Vote | - | $56.67 | - | $155.17 | $211.84 |
| | Adjudication | Gemini 1.5 Pro | 3.5 | $263.49 | 10.5 | $389.65 | $653.15 |
| Intervention | Question Answering | GPT-4o Mini | 0.15 | $4.25 | 0.6 | $12.56 | $16.81 |
| | | Gemini 1.5 Flash | 0.075 | $2.12 | 0.3 | $5.55 | $7.67 |
| | | Claude 3 Haiku | 0.25 | $7.08 | 1.25 | $34.65 | $41.73 |
| | Majority Voting | Soft-Vote | - | $13.45 | - | $52.76 | $66.21 |
| | Debating | GPT-4o Mini | 0.15 | $28.77 | 0.6 | $68.74 | $97.51 |
| | | Gemini 1.5 Flash | 0.075 | $21.11 | 0.3 | $61.76 | $82.88 |
| | | Claude 3 Haiku | 0.25 | $38.98 | 1.25 | $94.45 | $133.43 |
| | Vorting on debating | MAD-Soft-Vote | - | $88.86 | - | $224.95 | $313.81 |
| | Adjudication | Gemini 1.5 Pro | 3.5 | $394.11 | 10.5 | $565.80 | $959.90 |
